# AutoRADP: An Interpretable Deep Learning Framework to Predict Rapid Progression for Alzheimer’s Disease and Related Dementias Using Electronic Health Records

**DOI:** 10.1101/2025.04.06.25325337

**Authors:** Qiang Yang, Weimin Meng, Pei Zhuang, Stephen Anton, Yonghui Wu, Rui Yin

**Affiliations:** Department of Health Outcomes and Biomedical Informatics, University of Florida; Department of Neurosurgery, Brigham and Women’s Hospital, Harvard Medical School; Department of Physiology and Aging, University of Florida

## Abstract

Alzheimer’s disease (AD) and AD-related dementias (ADRD) exhibit heterogeneous progression rates, with rapid progression (RP) posing significant challenges for timely intervention and treatment. The increasingly available patient-centered electronic health records (EHRs) have made it possible to develop advanced machine learning models for risk prediction of disease progression by leveraging comprehensive clinical, demographic, and laboratory data. In this study, we propose AutoRADP, an interpretable autoencoder-based framework that predicts rapid AD/ADRD progression using both structured and unstructured EHR data from UFHealth. AutoRADP incorporates a rule-based natural language processing method to extract critical cognitive assessments from clinical notes, combined with feature selection techniques to identify essential structured EHR features. To address the data imbalance issue, we implement a hybrid sampling strategy that combines similarity-based and clustering-based upsampling. Additionally, by utilizing SHapley Additive exPlanations (SHAP) values, we provide interpretable predictions, shedding light on the key factors driving the rapid progression of AD/ADRD. We demonstrate that AutoRADP outperforms existing methods, highlighting the potential of our framework to advance precision medicine by enabling accurate and interpretable predictions of rapid AD/ADRD progression, and thereby supporting improved clinical decision-making and personalized interventions.

## Introduction

Alzheimer’s disease (AD) and AD-related dementias (ADRD) represent a significant global health challenge, affecting over 40 million individuals worldwide, and the number will increase to more than 100 million by 2050^1^. These progressive neurodegenerative disorders are characterized by heterogeneous progression rates, with some patients experiencing a gradual decline over decades, while others exhibit rapid progression (RP), marked by swift and severe deterioration in cognitive, functional, and behavioral abilities within a few months or years^2,3^. Patients with RP experience accelerated deterioration in memory, executive function, and daily living skills, leading to a faster transition to severe stages of the disease compared to those with slower progression^2,3^. This rapid decline is often accompanied by pronounced behavioral and psychological symptoms, such as agitation, aggression, and mood disturbances, which further complicate care and management^4^. The outcomes of RP are particularly devastating, as patients require more intensive caregiving support, experience a reduced quality of life, and face higher mortality rates^5^. In clinical practice, not all individuals with AD progress to severe stages of the disease, and predicting who will progress rapidly remains challenging^6^. To assess disease severity and track progression, cognitive assessments, such as the Mini-Mental State Examination (MMSE), Montreal Cognitive Assessment (MoCA), and Clinical Dementia Rating (CDR)^7^, etc., are widely used. For example, Kinkingn éhun et al. defined the RP as a decrease of ≥ 6 points on the MMSE over 36 months^8^, while Tan et al. proposed ≥ 2 points on the MoCA over 6 months^9^.

The advancement of machine learning (ML) has revolutionized disease risk prediction, particularly through the use of electronic health records (EHRs)^10,11^, which offer a rich repository of data, including demographic information, clinical assessments, and laboratory results. For example, Akter et al. utilized the traditional machine methods such as Gradient-Boosted Trees (GBT), Light Gradient-Boosting Machine (LightGBM), Random Forest (RF), for the early diagnosis and prediction of ADRD using structured EHRs from MU Healthcare^12^. Fisher et al. introduced a Conditional Restricted Boltzmann Machine to simulate detailed, personalized disease progression trajectories for patients with MCI or AD using cognitive assessment and laboratory tests data from the Coalition Against Major Diseases Online Data Repository for AD^13^. Furthermore, Meng et al. proposed an interpretable population graph network framework for identifying RP from MCI to AD by utilizing structured EHRs from UKBioBank^14^. Ma et al. built deep learning models using recurrent neural networks with attention mechanisms to predict RP using data extracted from EHRs^15^. However, these approaches often neglect unstructured data, such as clinical notes, containing valuable contextual information and patient observations, which is critical to trace the progression rate for AD/ADRD patients.

In this study, we propose AutoRADP, a novel interpretable **<u>auto</u>**encoder-based framework to predict **<u>r</u>**apid **<u>AD</u>**/ADRD **<u>p</u>**rogression leveraging both unstructured and structured EHR data from UF Health. The overall workflow of our proposed framework is illustrated in **Figure 1**. Specifically, we used a rule-based natural language process (NLP) method following Chen et al.^16^ to extract critical AD/ADRD patient outcomes (e.g., MMSE and MoCA scores) from clinical notes, which were further employed to label the RP of AD/ADRD patients using the decline rate per year^17,18^. We also leveraged feature selection techniques to derive important routine EHR information, e.g., demographics, laboratory results, and treatments. To address data imbalance issue, we introduced a hybrid sampling method that combines similarity-based and clustering-based upsampling, ensuring robust model training. Then, an autoencoder-based classifier, consisting of the autoencoder layer and the Multi-Layer Perceptron (MLP) layer, was designed for the RP prediction of AD/ADRD patients. Additionally, we leveraged SHapley Additive exPlanations (SHAP) values to interpret the importance of features for the model’s predictions. The extensive experiments demonstrated our proposed AutoRADP can achieve superior performance to identify RP for AD/ADRD patients with interpretability, making it a valuable tool for clinicians and researchers in advancing precision medicine.

**Figure 1:**
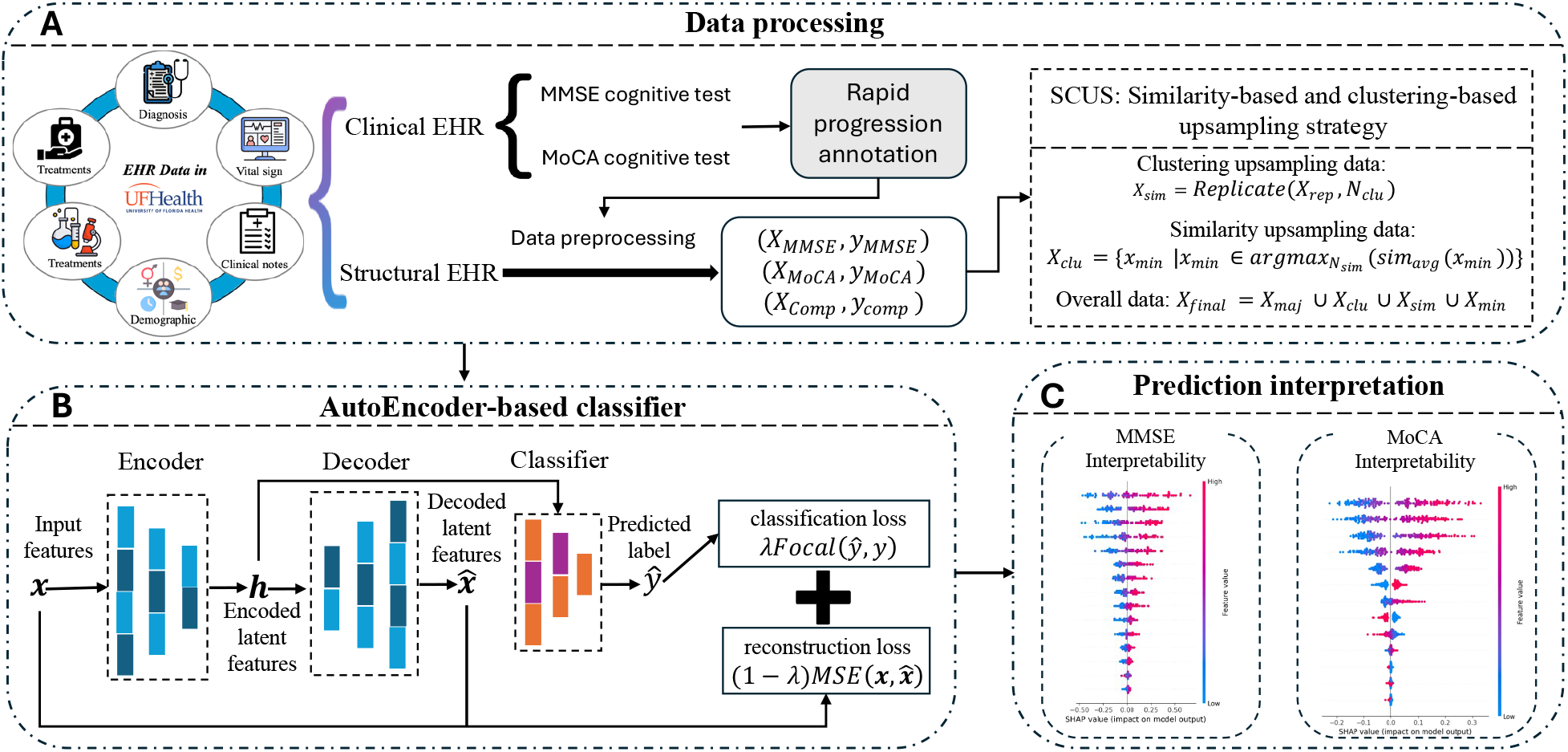
Illustration of the proposed framework AutoRADP. (A) Data preprocessing and extraction from structured and unstructured EHRs, along with sample enrichment. (B) The autoencoder-based classifier for learning the representations and identifying RPrs of AD/ADRD. (C) SHAP values to interpret the predictions through the feature importances.

## Materials and Methods

### Problem formulation

To identify RP of AD/ADRD using EHR data, we utilized the variation of cognitive assessments (i.e., MMSE, and MoCA scores) to measure the progression rate of AD/ADRD, which can be extracted from the clinical notes. It is notable that for each cognitive assessment, a patient would have multiple records. Using MMSE as an example, we defined the earliest available MMSE score as the baseline score *score*_*base*_, and the associated date as the index date. We then selected the nearest MMSE assessment occurring at least six months after the index date as the target score *score*_*target*_, and its corresponding date as the target date. We calculated the progression rate 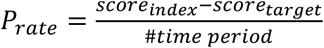 Following established practices in the literature^17,18^, we defined rapid progression (RP) based on the following conditions: *P*_*rate*_ ≥ 3 for AD/ADRD patients using MMSE assessment. If this criterion was satisfied, we labeled these AD/ADRD patients as RPrs, otherwise, non-RPrs, which are referred to as positive and negative cases. Similarly, we will classify cognitive change greater than 3 points/year as rapid progression for AD/ADRD patients using MoCA assessment^9^.

### Dataset construction

This study utilized Electronic Health Records (EHRs), including clinical notes and structured records, from the UF Health Integrated Data Repository (IDR), which aggregates data from various clinical and administrative systems. Patients diagnosed with AD/ADRD between January 1, 2012, and December 31, 2019, were included in the study. For the ADRD, we focused on several common ones, including vascular dementia, Lewy body dementia, and frontotemporal dementia. The diagnostic criteria were based on ICD codes following: Alzheimer’s disease (ICD-9: 331.0; ICD-10: G30, G30.0, G30.1, G30.8, G30.9), vascular dementia (ICD-9: 290.4, 290.40, 290.41, 290.42, 290.43; ICD-10: F01, F01.5, F01.50, F01.51), Lewy body dementia (ICD-9: 331.82; ICD-10: G31.83), and frontotemporal dementia (ICD-9: 331.1, 331.11, 331.19; ICD-10: G31.0, G31.01, G31.09). This study was approved by the University of Florida Institutional Review Board (IRB) under protocol no. IRB202001888.

To extract MMSE and MoCA scores from clinical notes, we employed a rule-based natural language processing (NLP) method following the approach by Chen et al.^16^. This resulted in 1,835 unique AD/ADRD patients with MMSE scores and 1,796 patients with MoCA scores, among which 300 patients had both types of assessments. To ensure data quality for evaluating cognitive changes, we applied the following exclusion criteria to each patient for each assessment type: (1) patients with only one assessment record were excluded; (2) for patients with two assessment records, only those whose second assessment occurred at least six months after the first were retained; and (3) for patients with more than two assessment records, the first assessment was used as the baseline, and the earliest follow-up assessment at least six months later was selected as the target. Based on these criteria, we constructed two primary groups: the MMSE group, which included 1,122 patients (202 rapid progressors [RPrs] and 920 non-rapid progressors [non-RPrs]), and the MoCA group, which included 1,001 patients (168 RPrs and 833 non-RPrs). Additionally, we created a composite group consisting of patients with both MMSE and MoCA scores. To address inconsistencies in RP classification between the two assessment types, we excluded cases with conflicting RP/non-RP labels. This resulted in a final composite group of 157 patients, comprising 20 RPrs and 137 non-RPrs.

### Feature preprocess and selection

The clinical information of selected patients included a wide range of features, which were categorized into continuous (e.g., body mass index, age) and categorical features (e.g., sex, diagnostic codes). Continuous features were normalized using Min-Max scaling to map values to a 0–1 range, ensuring comparability across features. Categorical features were transformed through one-hot encoding. We then removed the features with missingness rate over 30% and applied four different feature selection methods, including variance threshold, Pearson Correlation, Chi2 test and ANOVA test, to further filter out the remaining features that were less relevant to our predictive targets by the following rules (1) Variance threshold < 0.1; (2) Pearson Correlation between features and targets < 0.01; (3) P-value ≥ 0.05 by Chi2 test; and (4) P-value ≥ 0.05 by ANOVA test. We ended up with 325 features for the MMSE group, and 127 features for the MoCA group. For the composite group, we first applied the selected feature names from MMSE group and MoCA group and extract AD/ADRD features with MMSE and MoCA scores. This means that the composite group had two types of features, i.e., features in MMSE group and MoCA group. To handle missing values in the selected features, we utilized K-nearest neighbors (k = 3) algorithm for imputation.

### Similarity-based and clustering-based upsampling strategy (SCUS)

Data imbalance is a common issue in machine learning tasks, often leading to biased models that disproportionately favor the majority class that results in suboptimal performance^19,20^. To address this challenge, various data sampling methods have been developed, including under-sampling, oversampling, and synthetic sampling^21–23^. However, these methods may fail to capture the complexity and heterogeneity inherent in EHRs, potentially producing unrealistic combinations of clinical features that do not accurately represent true patient profiles^24,25^. To address these limitations, we proposed SCUS (Similarity-based and Clustering-based Upsampling Strategy), a hybrid upsampling method that combines two complementary approaches to balance datasets while preserving the diversity and integrity of the data. SCUS generates additional samples from the minority class to achieve a desired positive-to-negative ratio.

#### Clustering-based Upsampling

In the clustering-based upsampling method, K-Means clustering was applied to the minority class samples (*X*_*min*_) to form *m* = min (*N*_*clu*_, |*X*_*min*_|) clusters where *N*_*clu*_ is the number of samples needed from clustering-based upsampling. For each cluster *j*, the centroid *c*_*j*_ was computed, and the sample closest to the centroid was selected as the candidates:

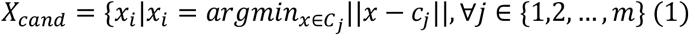

Here, *C*_*j*_ is the set of samples belonging to the cluster *j*. If the number of candidates |*X*_*cand*_| was less than *N*_*clu*_, we replicated the samples in *X*_*rep*_to meet the required count, i.e., *N*_*clu*_− |*X*_*rep*_|, with the equation *X*_*clu*_,*= Replicate*(*X*_*rep*_, *N*_*clu*_). Otherwise, we used the first *N*_*clu*_ samples: *X*_*clu*_ *= X*_*rep*_[: *N*_*clu*_]. This method ensures that we obtain the desired number of samples while maintaining the diversity and representativeness of the minority class.

#### Similarity-Based Upsampling

For the similarity-based upsampling method, we calculated the cosine similarity between each minority sample and the majority class samples.

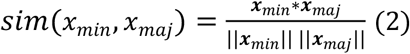

where *x*_*maj*_ ∈ *X*_*maj*_ and *X*_*maj*_ is the majority class samples. ***x***_*min*_ and ***x***_*maj*_ represented the features of data samples *x*_*maj*_ and *x*_*maj*_, respectively. For each minority sample *x*_*min*_, the average similarity to all majority samples was computed:

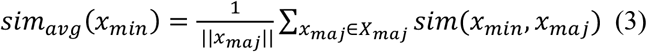

The minority samples with the highest average similarity scores were prioritized for duplication:

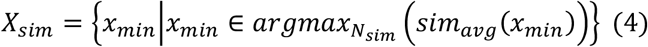

#### Weighted Combination

The final hybrid upsampled dataset combined the samples generated from the clustering-based and similarity-based methods where the total number of minority samples required was *N*_*desired*_*= pnr* · |*X*_*maj*_|. Given the weights *w*_*clu*_ and *w*_*sim*_ of the clustering-based and similarity-based data samples such that *w*_*clu*_ + *w*_*sim*_ *=* 1, the number of samples generated from each method was *N*_*clu*_ *= w*_*clu*_ · *N*_*desired*_ and *N*_*sim*_ = *w*_*sim*_ · *N*_*desired*_. The final oversampled dataset combined the results from both clustering-based upsampling samples (*X*_*clu*_) and similarity-based upsampling samples (*X*_*sim*_): *X*_*final*_ *= X*_*maj*_ ∪ *X*_*clu*_ ∪ *X*_*sim*_ ∪ *X*_*min*_. The corresponding labels were also combined to form the balanced dataset.

### Autoencoder-based classifier

*Autoencoder for Feature Extraction*. The autoencoder is designed to compress the input data into a lower-dimensional latent space through an encoder-decoder structure. The encoder progressively reduces the input dimensionality through a series of fully connected layers with ReLU activation functions. The decoder then reconstructs the input from the latent representation using a symmetric architecture. The encoder (*f*_*enc*_) and decoder (*f*_*dec*_) can be described as follows, where ***x*** is the input features, ***h*** is the latent features, and 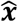 is the reconstructed input features. *W*_*i*_ and *b*_*i*_ are the learnable parameters. This process helps in capturing the essential features of the data while discarding noise.

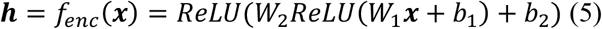

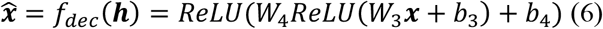

#### MLP for Classification

The classifier operates on the latent representation ***h*** produced by the encoder. It consists of a fully connected layer followed by batch normalization, and ReLU activation for regularization. The final layer outputs a single value (logit), which can be passed through a sigmoid function for binary classification. The classifier can be expressed below. Here, *y* is the predicted output, *W*_*i*_ and *b*_*i*_ are the learnable parameters.

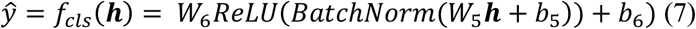

#### Model Training

The training process for the autoencoder-based classifier involves optimizing a combined loss function that integrates both classification loss and reconstruction loss. The classification loss was implemented using the Focal Loss (*L*_*cls*_), which is particularly effective for addressing class imbalance by focusing on hard-to-classify samples and down-weighting the loss for well-classified samples. It is formulated below:

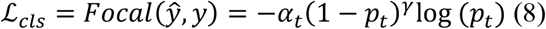

Here, *y* is the ground truth label. *p*_*t*_ is the predicted probability for the true class, which is obtained by applying a sigmoid function to 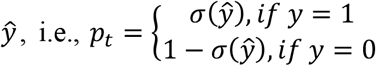, where 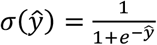 is the sigmoid function. For reconstruction, we used the Mean Squared Error (MSE) Loss, which measures the difference between the original input and the reconstructed input as:

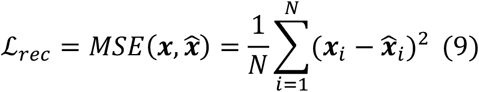

where N is the number of samples, and **x** and 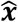 are the original and reconstructed features, respectively. The total loss is a weighted combination of the two losses:

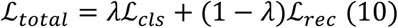

where λ is a weighting parameter that controls the trade-off between classification and reconstruction.

#### Ensemble Model for RP Prediction of AD/ADRD

After training the MMSE-based AutoRADP and MoCA-based AutoRADP models on the MMSE and MoCA groups, respectively, we constructed an ensemble model to enhance predictive performance on the composite group, which includes patients with both MMSE and MoCA assessments. For each patient in this group, we obtained the predicted probabilities from both the MMSE-based and MoCA-based models. These probabilities were then concatenated to form integrated feature vectors, capturing complementary predictive signals from the two modalities. We subsequently trained a Logistic Regression classifier on these integrated features to produce the final RP predictions. The output probabilities were compared against a predefined threshold to evaluate the ensemble model’s classification performance.

### Model interpretation

Understanding how a model makes predictions is as crucial as its predictive performance, particularly in biomedical domains where interpretability fosters trust and enables insights. To achieve this, we designed the SHAP^26^-based interpreters on the trained MMSE-based model and MoCA-based models, respectively.

Specifically, AD/ADRD features with MMSE scores from the composite group, along with the trained model on the MMSE group, was used to initialize a SHAP interpreter. The corresponding SHAP values were then calculated using DeepSHAP, which quantify the contribution of each feature to the model’s predictions. These values provide interpretable insights into the importance of individual features. Similarly, we applied the same SHAP-based interpretation approach for AD/ADRD features with MoCA scores from the composite group. To check the consistence of the interpretation, we tried to compare the top ranked features for the same ones. This approach not only enhances transparency but also helps clinicians and researchers identify key factors driving RP in AD/ADRD.

### Experimental setup

#### Implementation and evaluation

Each group-specific dataset was randomly divided into training (70%), validation (10%), and testing (20%) sets. Rapid progressors (RPrs) were labeled as “1” (positive class), while non-rapid progressors (Non-RPrs) were labeled as “0” (negative class). The AutoRADP model was trained for 80 epochs using a batch size of 16 and a learning rate of 0.001. To mitigate overfitting, we applied a dropout rate of 0.6 in the MLP layers and implemented early stopping, which halted training if the validation performance failed to improve over 20 consecutive epochs. Model training was conducted on the training set, with performance subsequently evaluated on the validation and testing sets. To further investigate the impact of data oversampling strategies, we explored four upsampling techniques: random-based, dissimilarity-based, similarity-based, and clustering-based methods. Random-based upsampling balances the dataset by randomly duplicating minority class samples to match the desired positive-to-negative ratio. The dissimilarity-based approach selects minority samples that are least similar to the majority class, in contrast to the similarity-based method, which chooses samples most similar to it. Clustering-based upsampling involves grouping minority class samples into clusters and resampling within each cluster to preserve diversity. For comparison, we also evaluated a baseline condition without any oversampling, called Default.

To ensure consistent and reproducible model performance across experiments, key parameter settings are detailed: the Focal loss parameters (*α* and *γ*) were set to 0.3 and 2.0 in the MMSE group and 0.5 and 3.0 for the MoCA group, respectively. The weights for clustering-based and similarity-based sampling were 8:2 for the MMSE group and 9:1 for the MoCA group, while the positive-to-negative ratio was set to 1.0 for both groups. Additionally, the balance weight (*λ*) in Equation (10), controlling the trade-off between classification and reconstruction loss, was set to 0.5 for both groups. These settings were carefully optimized to ensure consistent and reproducible model performance across experiments. We employed four distinct metrics to evaluate the prediction, including sensitivity, F1-score, the area under the receiver operating characteristic curve (AUROC), and the area under the precision-recall curve (AUPRC).

#### Benchmark approaches

We benchmarked our designed AutoRADP against several baseline models across two categories: traditional machine learning (ML) methods, and existing deep learning (DL) approaches for rapid progression of AD/ADRD diagnosis. For traditional ML models, we utilized gaussian naive bayes (GNB), stochastic gradient descent (SGD), support vector machine (SVM), and random forest (RF). Additionally, we compared our model with advanced DL architectures, including convolutional neural networks^27^ (CNN), multi-layer perceptron^28^ (MLP), and multi-head attention(MHAttn)^29^.

## Results

### Descriptive statistics of selected AD/ADRD patients

The descriptive statistics of the patient data, as summarized in **Table 1**, provides a snapshot of the study group’s demographic, clinical, and laboratory characteristics. In both the MMSE (N=1,122) and MoCA (N=1,001) groups, according to the table, we can find that the age of RPrs of AD/ADRD patients in both groups were elder, with mean ages of 76.3 ± 9.5 (MMSE group) and 75.8 ± 9.3 (MoCA group), compared to 75.1 ± 9.1 and 73.8 ± 9.3 in the non-RPrs groups. In the MMSE group, 61.0% of the RPrs were female, slightly higher than the non-RPrs (58.0%), whereas in the MoCA group, 51.0% of the RPrs were female and 54.0% of them are non-RPrs. A higher proportion of White participants was observed in the non-RPrs (84.8% in both group) compared to 75.7% (RPrs in MMSE group) and 83.9% (RPrs in MoCA group). Additionally, we observed that HbA1c levels were notably elevated in the RPrs of the MoCA group (8.1 ± 3.1%) versus 6.6 ± 1.8% in the non-RPrs, while RPrs patients in the MMSE group had a slightly lower HbA1c (6.4 ± 0.7%) compared to non-RPrs (6.7 ± 1.9%). Glucose levels were higher in the non-RPrs (129.3 ± 52.0 mg/dL in MMSE group and 133.4 ± 61.5 mg/dL in MoCA group) than in the RPrs (126.2 ± 51.4 mg/dL and 120.5 ± 36.6 mg/dL, respectively). For the medication use of these patients, it was suggested that Donepezil and Memantine were more frequently prescribed in the RPrs(73.8% vs. 55.4% and 54.0% vs. 38.6% in the MMSE group; 63.1% vs. 47.3% and 39.9% vs. 30.1% in the MoCA group), while statin use remained similar in RPrs and non-RPrs across different groups. Interesting, most patients indicated no history of smoking (over 97% on average), and there are no significant differences on BMI and cholesterol levels across RPrs and non-RPrs across different groups.

**Table 1.** Descriptive statistics on the characteristics of the study cohort.

|  | MMSE group (N=1,122) |  | MoCA group (N=1,001) |  |
| --- | --- | --- | --- | --- |
|  | RPrs (202) | Non-RPrs (920) | RPrs (168) | Non-RPrs (833) |
| **Demographics** |  |  |  |  |
| Age | 76.3 (9.5) | 75.1 (9.1) | 75.8 (9.3) | 73.8 (9.3) |
| Sex |  |  |  |  |
| Female, N (%) | 123 (61.0%) | 532 (58.0%) | 86 (51.0%) | 452 (54.0%) |
| Male, N (%) | 79 (39.0%) | 388 (42.0%) | 82 (49.0%) | 381 (46.0%) |
| **Ethnicity, N (%)** |  |  |  |  |
| Non-Hispanic | 11 (5.4%) | 88 (9.6%) | 17 (10.1%) | 95 (11.4%) |
| Unknown | 191 (94.6%) | 832 (90.4%) | 151 (89.9%) | 738 (88.6%) |
| **Race, N (%)** |  |  |  |  |
| White | 153 (75.7%) | 706 (84.8%) | 141 (83.9%) | 706 (84.8%) |
| Unknown | 49 (24.3%) | 127 (15.2%) | 27 (16.1%) | 127 (15.2%) |
| **Smoking status, N (%)** |  |  |  |  |
| Never | 196 (97.0%) | 886 (96.3%) | 165 (98.2%) | 809 (97.1%) |
| Unknown | 6 (3.0%) | 34 (3.7%) | 3 (1.8%) | 24 (2.9%) |
| **Laboratory measures** |  |  |  |  |
| BMI, kg/m<sup>2</sup> | 26.4 (5.5) | 26.8 (5.8) | 26.6 (5.2) | 27.5 (5.8) |
| HbA1c, % | 6.4 (0.7) | 6.7 (1.9) | 8.1 (3.1) | 6.6 (1.8) |
| Cholesterol, mg/dL | 171.6 (46.2) | 174.6 (43.0) | 172.5 (39.2) | 170.4 (44.7) |
| HDL, mg/dL | 57.3 (18.6) | 56.4 (20.2) | 55.1 (18.7) | 55.1 (20.1) |
| LDL, mg/dL | 91.1 (36.6) | 95.6 (45.3) | 95.5 (42.2) | 93.7 (48.1) |
| Glucose, mg/dL | 126.2 (51.4) | 129.3 (52.0) | 120.5 (36.6) | 133.4 (61.5) |
| **Medications** |  |  |  |  |
| Pravastatin | 35 (17.3%) | 197 (21.4%) | 28 (16.7%) | 161 (19.3%) |
| Simvastatin | 54 (26.7%) | 247 (26.8%) | 42 (25.0%) | 199 (23.9%) |
| Donepezil | 149 (73.8%) | 510 (55.4%) | 106 (63.1%) | 394 (47.3%) |
| Memantine | 109 (54.0%) | 355 (38.6%) | 67 (39.9%) | 251 (30.1%) |
| Namenda | 17 (8.4%) | 81 (8.8%) | 14 (8.3%) | 45 (5.4%) |

Continuous variables were presented as “mean (std)”; categorical variables were presented as “number (percentage %)”

### Comparative performance of our model against benchmark approaches

**Table 2** presents the performance comparison between the proposed AutoRADP framework and various baseline classifiers for identifying rapid progression (RP) in AD/ADRD patients within the MMSE and MoCA groups. Among the deep learning models, the Multi-Layer Perceptron (MLP) demonstrated moderate sensitivity and F1 scores but underperformed in terms of AUROC and AUPRC. CNN performed slightly better, achieving the highest AUPRC in the MMSE group and the highest AUROC in the MoCA group, suggesting its effectiveness in capturing complex feature interactions and non-linear patterns. MHAttn model showed competitive sensitivity in the MoCA group but yielded relatively lower AUROC and AUPRC scores. Among traditional machine learning benchmarks, GNB achieved the highest AUROC in the MMSE group but had poor F1 and AUPRC scores, indicating its limitations in handling class imbalance. SGD and SVM models showed moderate performance, with SVM achieving strong AUROC in the MoCA group but performing poorly in sensitivity and F1 score. RF displayed the weakest results across all metrics, indicating its unsuitability for this prediction task. In contrast, the proposed AutoRADP model consistently outperformed all baselines in both sensitivity and F1 score across groups. Although it did not achieve the highest AUROC and AUPRC, its values remained competitive, reflecting a balanced performance. Additionally, across all models, higher sensitivity and F1 scores were observed in the MMSE group compared to the MoCA group, which may be attributed to differences in feature sets used during model training. This suggests that the features derived from the MMSE group may be more discriminative for identifying rapid progression. On the other hand, models trained on the MoCA group showed relatively better AUROC and AUPRC values, with CNN achieving the highest scores on both metrics.

### Ensemble strategy to improve predictive performance

To enhance the identification of rapid progressors (RPrs), we investigated an ensemble strategy that combines predictions from the MMSE- and MoCA-based AutoRADP models. This approach was evaluated on the composite group using a logistic regression model, where the input features consisted of the predicted probabilities of RPrs classification from each individual model. The ensemble model demonstrated improved predictive performance, achieving a sensitivity of 0.900—surpassing the sensitivities of 0.819 and 0.810 achieved by the MMSE-based and MoCA-based models, respectively. These findings indicate that integrating predictions from multiple cognitive assessment-based models can effectively enhance the detection of rapid progressors (RPrs).

**Table 2.**
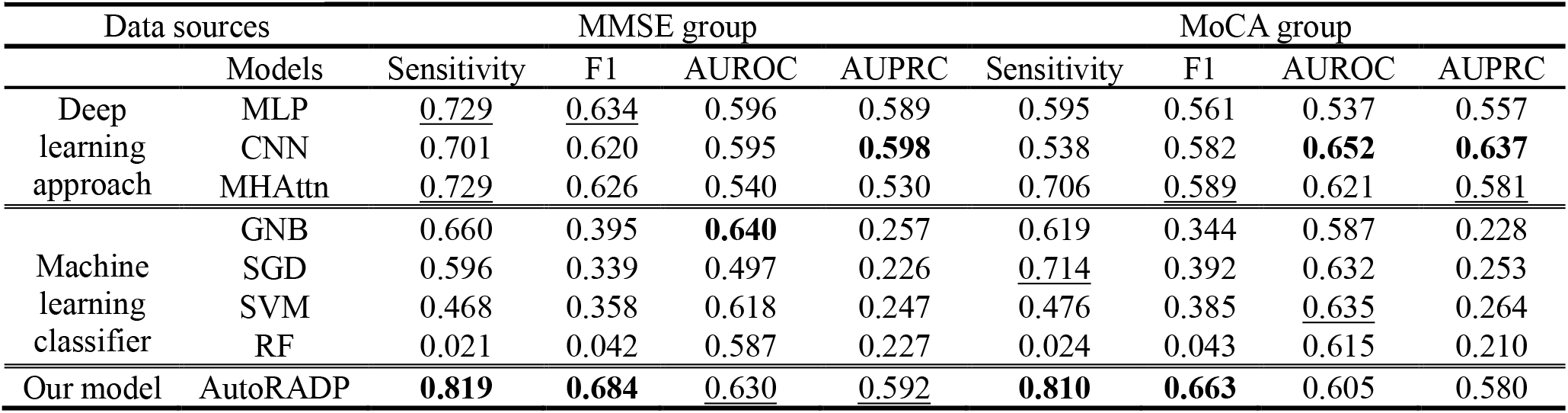
Performance comparison between our proposed framework and baseline classifiers on the MMSE group and MoCA group. The best-performing models in each metric are marked in bold, while the second-best are indicated with underlining.

| Data sources |  | MMSE group |  |  |  | MoCA group |  |  |  |
| --- | --- | --- | --- | --- | --- | --- | --- | --- | --- |
|  | Models | Sensitivity | F1 | AUROC | AUPRC | Sensitivity | F1 | AUROC | AUPRC |
| Deep learning approach | MLP | <u>0.729</u> | <u>0.634</u> | 0.596 | 0.589 | 0.595 | 0.561 | 0.537 | 0.557 |
|  | CNN | 0.701 | 0.620 | 0.595 | **0.598** | 0.538 | 0.582 | **0.652** | **0.637** |
|  | MHAttn | <u>0.729</u> | 0.626 | 0.540 | 0.530 | 0.706 | <u>0.589</u> | 0.621 | <u>0.581</u> |
| Machine learning classifier | GNB | 0.660 | 0.395 | **0.640** | 0.257 | 0.619 | 0.344 | 0.587 | 0.228 |
|  | SGD | 0.596 | 0.339 | 0.497 | 0.226 | <u>0.714</u> | 0.392 | 0.632 | 0.253 |
|  | SVM | 0.468 | 0.358 | 0.618 | 0.247 | 0.476 | 0.385 | <u>0.635</u> | 0.264 |
|  | RF | 0.021 | 0.042 | 0.587 | 0.227 | 0.024 | 0.043 | 0.615 | 0.210 |
| Our model | AutoRADP | **0.819** | **0.684** | <u>0.630</u> | <u>0.592</u> | **0.810** | **0.663** | 0.605 | 0.580 |

### Performance evaluation under different oversampling strategies

**Table 3** highlights the superior performance of the SCUS method across both the MMSE and MoCA groups for AD/ADRD rapid progression. In the MMSE group, SCUS achieved the highest sensitivity (0.819) and F1 score (0.684) based on SCUS strategy for upsampling of the cohort. In contrast, traditional methods like Default and Clustering strategies performed significantly worse across these metrics. The Random strategy excelled in AUROC (0.676) and AUPRC (0.646) while securing the second-best sensitivity (0.723) and F1 score (0.668). For the MoCA group, SCUS also presented the highest sensitivity (0.810), F1 score (0.663), and AUPRC (0.580), along with a comparable AUROC (0.605). The Similarity strategy showed competitive performance with sensitivity (0.684), F1 score (0.600), and AUPRC (0.569) but was still worse than SCUS in overall effectiveness. When comparing performance across the MMSE and MoCA groups, our SCUS achieved comparable sensitivity and F1 scores across different upsampling methods in the MoCA group relative to the MMSE group. This suggests that models trained on the MoCA group may be equally effective in identifying rapid progression of AD/ADRD patients, despite differences in the cognitive domains measured by the assessments. However, we observed higher AUROC and AUPRC values in the MMSE group for most upsampling strategies, indicating that models trained on the MMSE group may offer better overall discriminative ability between rapid and non-rapid progressors.

### Model interpretation

We interpreted the predictions of the AutoRADP model using SHAP values to visualize feature importance within the composite group, as illustrated in **Figure 2**. In MMSE group (**Figure 2A**), the top positive contributors to the model’s prediction were related to donepezil, including terms such as “donepezil”, “donepezil 10 MG Oral Tablet”, and “donepezil 5 MG Oral Tablet”. Conversely, in MMSE group, “phenylephrine hydrochloride” and “80 ACTUAAT albuterol 0.09 MG/ACTUAAT Metered Dose Inhaler” were negatively associated with rapid progression, as indicated by their SHAP values. Additionally, “donepezil hydrochloride” and “memantine hydrochloride” showed a moderate influence on the model’s predictions. In the MoCA group (**Figure 2B**), “memantine” had the highest positive SHAP value, followed by “quetiapine 25 MG Oral Tablet” and “Diagnostic ultrasound of heart (echocardiogram)”, suggesting these features were most predictive of rapid progression in that group. In contrast, “hydrocodone” and “promethazine” exhibited negative SHAP values, indicating a negative contribution to the prediction of rapid progression. When comparing feature importance across the two groups (**Figure 2C**), certain features demonstrated distinct patterns. For example, “Hydroxyzine”, and “Sugammadex sodium” had a stronger influence in the MoCA group, while “Hydromorphone 2 MG/ML injectable solution” and “Heparin sodium, porcine 1000 UNT/ML prefilled syringe” were more impactful in the MMSE group. Additionally, some features, such as “1 ML morphine sulfate 2 MG/ML cartridge”, showed variable influence across both groups.

**Table 3.**
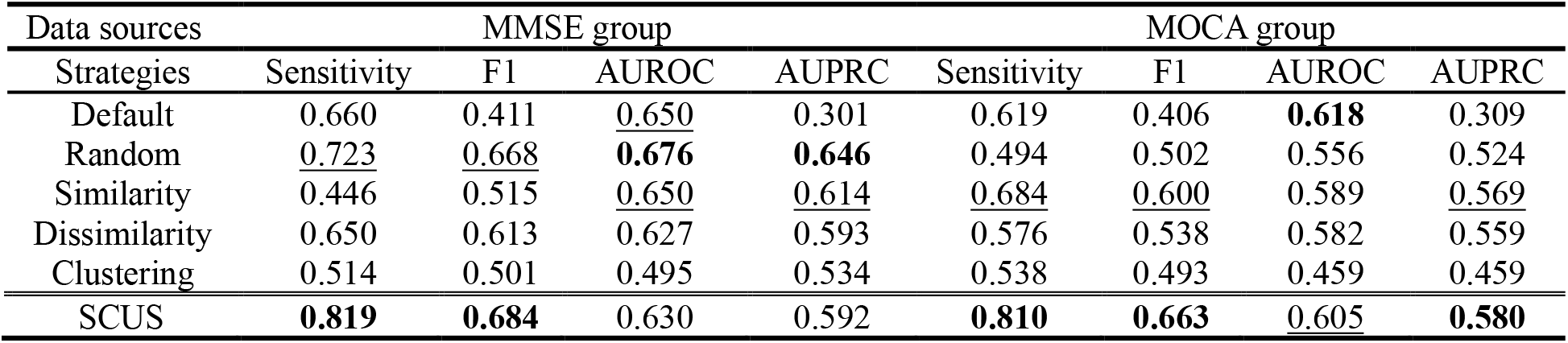
Performance comparison of different upsampling strategies on MMSE-based group and only MoCA-based group. The best-performing models in each metric are marked in bold, while the second-best are indicated with underlining.

**Figure 2:**
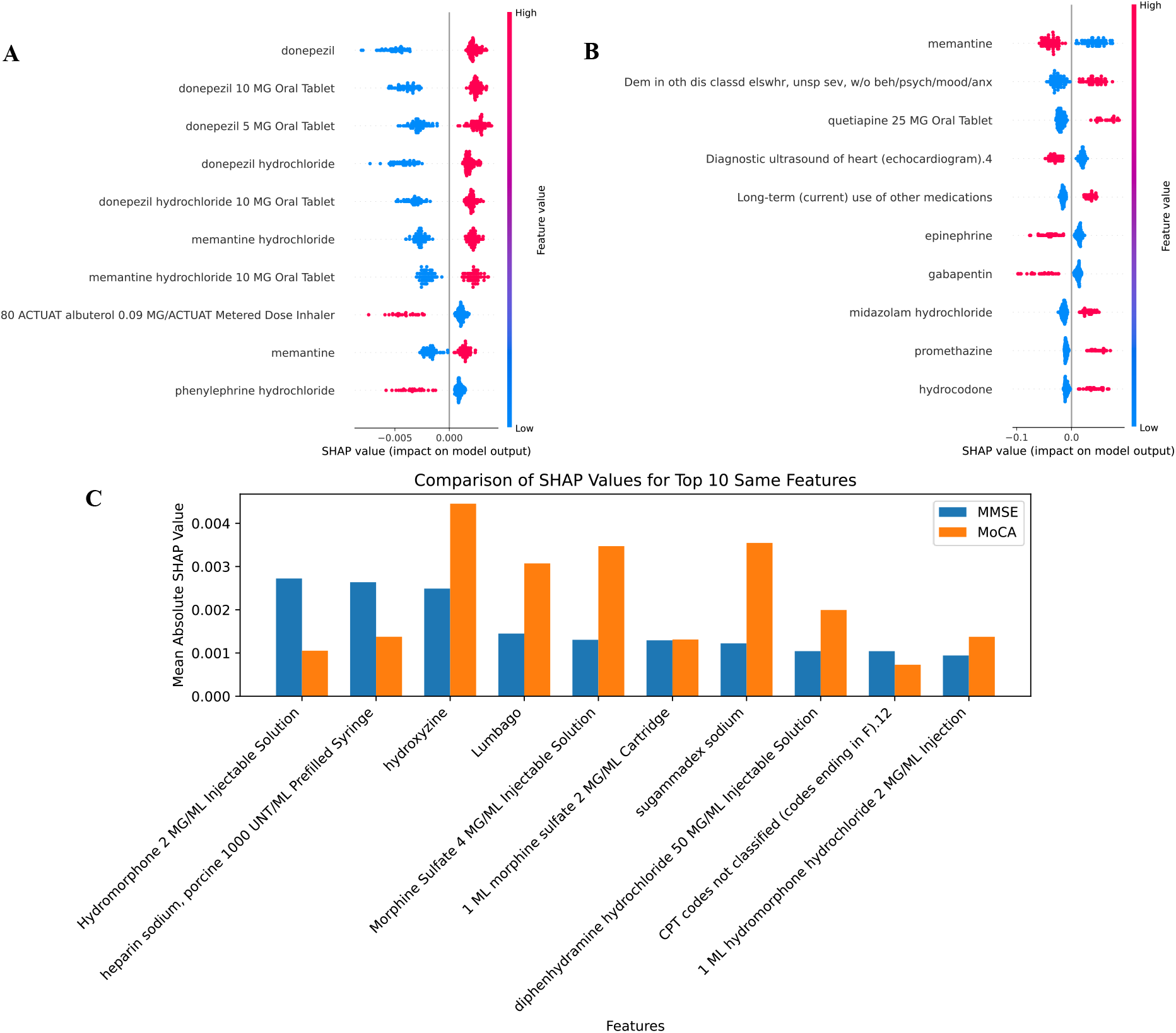
Feature importance visualization using SHAP values on the composite group. (A) Top 10 most important features identified in the MMSE group. (B) Top 10 most important features identified in the MoCA group. (C) Comparison of shared top 10 features across both groups.

## Discussion

This study developed and validated AutoRADP, an interpretable deep learning framework for predicting rapid progression (RP) for AD/ADRD patients using both unstructured and structured EHRs. On one hand, the unstructured EHRs, i.e., clinical notes, were used to extract the cognitive assessments, e.g., MMSE and MoCA scores, based on a rule-based NLP method. These scores were further processed to annotate the label of AD/ADRD patients (rapid progression vs. non-rapid progression) according to the cognitive decline per year. On the other hand, the structured EHRs involve diverse patient-centered information in different progression patterns, such as demographics, diagnoses, laboratory results, and treatments. To solve the data imbalance issue existing in our samples where rapid progressors are underrepresented, we proposed a hybrid sampling method, which leverages the benefits of similarity-based and clustering-based upsampling strategies to enable a relatively balanced distribution for model construction. An autoencoder-based classifier was designed to predict rapid progression of AD/ADRD using the selected features and annotated labels. To enhance the interpretation of the predictions of our proposed model, we applied the SHAP interpreter to identifying which features are the major contributors for the predictions. It is notable that our framework is not limited to only MMSE and MoCA scores, which is flexible to extend other cognitive assessments, such as Clinical Dementia Rating (CDR), Functional Activities Questionnaire (FAQ), Global Deterioration Scale (GDS), and Mini-Cog. The results show that AutoRADP effectively predicts RP of AD/ADRD patients with high sensitivity (0.819 in the MMSE group, 0.810 in the MoCA group, and 0.900 in the composite group), outperforming other baseline models. Additionally, SHAP-based model interpretation provided insights into critical features influencing RP predictions, with medications such as Donepezil and Memantine playing a crucial role.

Our results align with previous research emphasizing the utility of machine learning for identifying rapid AD/ADRD progression, including those developed by Meng et al.^14^ and Ma et al.^15^, which leveraged structured EHRs and deep learning techniques for RP identification. Nevertheless, AutoRADP distinguishes itself by (1) integrating both the structured and unstructured EHRs, where structured EHRs reflect the clinical characteristics and outcomes for these patients, and unstructured EHR (i.e., clinical notes) contain cognitive assessments (e.g., MMSE and MoCA) that can be used to measure the progression rate of AD/ADRD; and (2) leveraging a hybrid sampling approach to balance data distributions and providing interpretable predictions through SHAP analysis. Compared to prior models, AutoRADP demonstrates improved sensitivity while maintaining interpretability, making it more applicable in clinical practices to support early identification of AD/ADRD patients who are likely at risk for RP. The identification of key predictors, such as medication usage, and specific laboratory values that are associated with rapid AD/ADRD progression, can help optimize patient management strategies and refine treatment plans. While the AutoRADP framework demonstrates significant advancements, several limitations should be acknowledged. First, although SCUS addresses the class imbalance issue, it did not reflect the true prevalence of rapid progressors in real-world clinical settings, which may impact the model’s performance on external datasets. Second, we relied solely on cognitive changes measured by MMSE and MoCA scores to label rapid progression in AD/ADRD patients, due to data availability within our cohort. However, incorporating additional cognitive assessments, such as the CDR and GDS, could provide a more comprehensive and nuanced understanding of rapid progression. Finally, important factors derived from neuroimaging and genetic data, both of which are known to influence AD/ADRD progression, were not included in this study, potentially limiting the completeness of the analysis. Incorporating these modalities in future research could offer a more holistic understanding of disease mechanisms and progression.

## Conclusion

In summary, AutoRADP represents a significant advancement in predicting rapid progression in AD/ADRD patients by integrating cognitive assessments extracted from clinical notes and structured EHRs within an interpretable deep learning framework. Leveraging deep learning, the model enhances the ability to identify patients at higher risk of accelerated cognitive decline. Moreover, the model’s interpretability adds clinical value by offering transparent insights into key predictive features. These findings hold meaningful implications for the field, paving the way toward personalized AD/ADRD management and tailored therapeutic strategies that account for the heterogeneous progression patterns observed among patients.

## Data Availability

The data is private, which will be not released to the public.

## Acknowledgement

We acknowledge the University of Florida Integrated Data Repository (IDR) and the UF Health Office of the Chief Data Officer for providing the analytic data set for this project. Additionally, the Research reported in this publication was supported by the National Center for Advancing Translational Sciences of the National Institutes of Health under University of Florida Clinical and Translational Science Awards UL1TR000064 and UL1TR001427.

